# Longitudinal trends in the endemic symptomatic burden of COVID-19: Insights from community-based participatory virological surveillance in the Netherlands, November 2020 - April 2025

**DOI:** 10.1101/2025.10.09.25337658

**Authors:** Gesa Carstens, Wanda Han, Luisa Timmermans, Susan van den Hof, Dirk Eggink, Albert Jan van Hoek

## Abstract

**Objectives:** To analyze changes over time in symptomatology and symptom burden of SARS-CoV-2 infected individuals.

**Methods:** Data from symptomatic individuals from Infectieradar, an online community-based, participatory surveillance platform in the Netherlands, were analyzed (November 2020 – April 2025). Weekly questionnaires collected self-reported data on SARS-CoV-2 testing, health score (scale: 0–100), medication use, and healthcare visits. Longitudinal trends were evaluated using logistic regression models adjusted for age, sex, and comorbidities.

**Results:** During 4.5 years, 18,600 participants reported a positive SARS-CoV-2 test at least once. Upper respiratory symptoms became more prominent over time, and loss of smell and taste less. The self-reported health score and the proportion of symptomatic SARS-CoV-2-positive individuals with a high number of symptoms (more than 10 symptoms) remained relatively stable over time (October 2022 - April 2025), with a health impact comparable to influenza virus infections (n = 768).

**Conclusions:** The findings suggest that the clinical presentation of COVID-19 has evolved toward upper respiratory symptoms. Despite shifting symptoms, the impact on reported well-being in this population was remarkably stable, without a significantly lower endemic symptomatic burden. The health impact of COVID-19 is more comparable to influenza than to other common respiratory infections.

**Highlights:**

- Symptom profiles of COVID-19 shifted towards upper respiratory symptoms
- Rates of high number of symptoms among COVID-19 cases remained stable over time
- Likewise, the reported health score of COVID-19 case stayed constant over time COVID-19 had a similar disease burden as influenza

## INTRODUCTION

In the transition from a global pandemic to endemic circulation, SARS-CoV-2 (the virus causing COVID-19) continues to evolve, presenting new challenges in understanding its impact on public health. Besides antigenic evolution related to immune escape, an often-discussed aspect of viral evolution is the balance between a virus’s virulence—the harm it causes to its host—and its transmissibility – how easily it can spread. From an evolutionary standpoint, it is often theorized that viruses may eventually become less virulent (1). One concept suggests a balance between viral transmissibility and virulence; while a more virulent strain may have higher transmissibility, excessive harm to the host can reduce the effective transmission period. However, this view oversimplifies the complex evolution of many pathogens, including SARS-CoV-2. As an example, for SARS-CoV-2, transmission often occurs before severe symptoms develop, allowing the virus to spread effectively without being less virulent (2). This highlights the complex interaction between a virus’s transmissibility and its virulence.

Another factor influencing SARS-CoV-2 evolution and especially the observed clinical representation of the infection is the evolving immune profile of the population. This profile is shaped by an increasing proportion of individuals who have acquired some level of immunity through previous SARS-CoV-2 infections or vaccinations. In the Netherlands, vaccination campaigns began in January 2021 for priority groups and expanded to the general population in June 2021 (3). As more people gain immunity, the risk of severe disease decreases, leading to a reduced risk of hospitalization and mortality (4–6), even as new variants emerge.

Monitoring SARS-CoV-2 evolution in post-pandemic years is important for understanding the clinical representation of new evolving variants. While extensive research has focused on severe COVID-19 outcomes, including hospitalization and mortality, there is a gap in understanding the disease burden in the general population, among individuals with milder infections.

To address this gap we analyzed data from the Dutch online participatory surveillance platform Infectieradar with over 38,000 participants from the general community (7,8). We included data on symptomatic SARS-CoV-2 positive individuals (self-test or PCR) from November 2020 to April 2025. Participants reported symptoms and rated their perceived health in weekly questionnaires, enabling us to explore changes in self-reported health and symptom patterns of SARS-CoV-2 over time. With our community-based population we cannot directly assess the intrinsic virulence of circulating SARS-CoV-2. Instead, we focus on measuring the endemic symptomatic disease burden in the general population, defined as the symptoms and health effects experienced by people in the general population who contract COVID-19 but do not require hospitalization. These cases make up the majority of SARS-CoV-2 infections in an endemic setting, and monitoring them provides insight into the ongoing public health impact of the virus.

## METHODS

### Data source

The design and methods of the participatory surveillance study Infectieradar have been described previously (9). We included data from Infectieradar from the start of the online platform in November 2020 until the end of April 2025. Upon registration participants completed a background questionnaire gathering medical history information (including medication use for pre-existing conditions) as well as sociodemographic data (such as age, sex, and education level). Through weekly reminders via email, participants are asked to complete a brief questionnaire on a weekly basis, indicating if they had experienced any of 22 symptoms, and if they had taken a SARS-CoV-2 self-test in the previous week. Those who reported symptoms are asked additional questions about the start date of their symptoms, perceived health, healthcare-seeking behavior, medication use and absenteeism. Participants who report symptoms in more than one consecutive questionnaire are asked whether the current symptoms belong to the same infection episode as the previous ones, to allow data collection of the same infection over more than one week. In October 2022 a self-sampling component was included in Infectieradar (8,10). This component includes the distribution of SARS-CoV-2 antigen self-tests for participants to use (at home) when symptomatic, and material for a nose- and throat sample (NTS) to be sent by post to our laboratory for sequencing or multiplex PCR testing (RespiFinder 2SMART, PathoFinder, the Netherlands). Participants who took part in this were asked to submit a NTS if they reported a positive SARS-CoV-2 self-test (no older than 2 days) for sequencing or, when reporting a negative self-test, a random subset were asked to submit a NTS for multiplex PCR. Furthermore, in October 2022 an additional question was added for those with symptoms regarding the perceived health score (specified as a value between 0, indicating worst health imaginable, and 100, indicating best health imaginable). In this manuscript we look for the full period when it comes to reported symptoms and compare the self-reported health score from October 2022 onwards.

### Inclusion/exclusion criteria

Participants who filled in only the background questionnaire without completing at least two weekly symptom questionnaires were excluded. In addition, as our study design did not include testing of participants without symptoms, we only included SARS-CoV-2 positive participants with at least one symptom.

### Statistical analysis

#### Health indicators

We examined trends in healthcare-seeking behavior, medication use, absenteeism and the number of symptoms over time. Logistic regression models were used to analyze healthcare-seeking behavior, absenteeism and medication use per month of infection using aggregated episode data. These models were adjusted for age group (<25, 25–39, 40–49, 50–64, and 65+ years), sex, and pre-existing comorbidities (allergy, diabetes, chronic lung disease, and cardiovascular disease), using November 2020, the first month of our study period, as the reference month. Thus, when most people did not have prior immunity due to vaccination or previous infections. Moreover, in November 2020 the circulation of SARS-CoV-2 in our study population was stable, and the amount of events for the different health indicators was high enough to draw meaningful conclusions. For the analysis of the number of symptoms reported during infection, we applied a Poisson regression model, to analyze possible changes per month of infection, adjusted for age group, sex, and pre-existing comorbidities.

#### Symptom analysis

We used a logistic regression model corrected for sex, age group and pre-existing co-morbidities to calculate per month for each symptom separately the odds ratio (OR) of experiencing that symptom using November 2020 as reference group. As participants could report symptoms for one infection episode in more than one weekly questionnaire, symptom presence per infection per participant were aggregated.

#### Self-reported health score

We analyzed the self-reported health score of participants per month. For this, we performed a generalized linear mixed model with log link function to model self-reported health during infection. We added a random intercept per participant, allowing each participant to have their own baseline level of self-reported health, capturing individual differences that are not explained by the fixed variables in the model. To account for changes in self-reported health during disease progression we added the time since start of the symptoms as natural spline with three knots to the model. Additionally, the model was adjusted for age group, sex, and pre-existing co-morbidities. We used the R package emmeans (12) to graphically show the predicted marginal mean of the self-reported health over the infection episode stratified by age group corrected.

#### Testing propensity

To examine potential reporting bias, where participants with a higher symptom burden might be more likely to test for SARS-CoV-2, we categorized all symptomatic participants (independent of the cause of their infection) into three groups based on their number of symptoms: low (0-4 symptoms), moderate (5-10 symptoms), and high (more than 10 symptoms). The 25th and 75th percentiles of the reported number of symptoms of the SARS-CoV-2 positive participants were used as cutoffs for these categories. We then used a logistic regression model to predict the probability of participants performing a self-test, adjusting for age group, sex, pre-existing comorbidities, days since symptom onset (modeled as a natural spline), and an interaction term between symptom group (based on number of symptoms) and the month of symptom reporting. Analyses were restricted to data collected from October 2022 onwards, when self-tests became widely available. To visualize testing trends, we calculated marginal means for each symptom group over time. Additionally, we hypothesized that participants who received a positive self-test result might report more symptoms, as they are aware of the underlying cause. To investigate this, we conducted a Poisson regression analysis to model the number of symptoms reported by SARS-CoV-2 positive participants who used a self-test compared to those who were diagnosed solely through PCR testing. The analysis was adjusted for month of infection, age group, sex, and presence of comorbidities. We then used estimated marginal means to visualize the results over time.

#### Weighted number of symptoms and health score

To account for differences in testing behavior over time and to see if self-testers are more likely to report symptoms as they know their source of infection, we stratified all symptomatic participants in our study by the categorized number of symptoms, regardless of the underlying cause of infection. For each group—low, moderate, and high symptom counts—we estimated the proportion of cases attributable to SARS-CoV-2 using the corresponding positivity rates (see **Supplemental Part 3** for an example). Credibility intervals for these estimates were calculated from 1,000 bootstrapped samples. Additionally, we conducted a similar analysis by stratifying participants according to their self-reported health scores (low: <38; moderate: 38–65; high:>65) instead of symptom count. Since health scores were introduced later in the study, this analysis was limited to data collected between October 2022 and April 2025.

Finally, to compare SARS-CoV-2 results with other respiratory pathogens, we repeated these analyses for participants who tested positive for rhinovirus, Influenza A and B, and seasonal coronaviruses (NL63, HKU1, OC43, and 229E).

## RESULTS

Between November 2020 and the end of April 2025 38,792 participants filled in at least two weekly questionnaires, leading to a total of 2,798,266 questionnaires (**Table 1**). Of those participants, 35,737 reported symptoms in at least one weekly questionnaire and 18,600 reported a positive self-test or PCR test at least once. In our study population, the majority of participants were female (59%), over 50 years old (59%) and had a higher education level (59%). Cardiovascular disease was the most frequently reported underlying condition in the study population, reported by 8.6% of all patients and accounting for 22,8 % of all reported conditions.

**Table 1.**
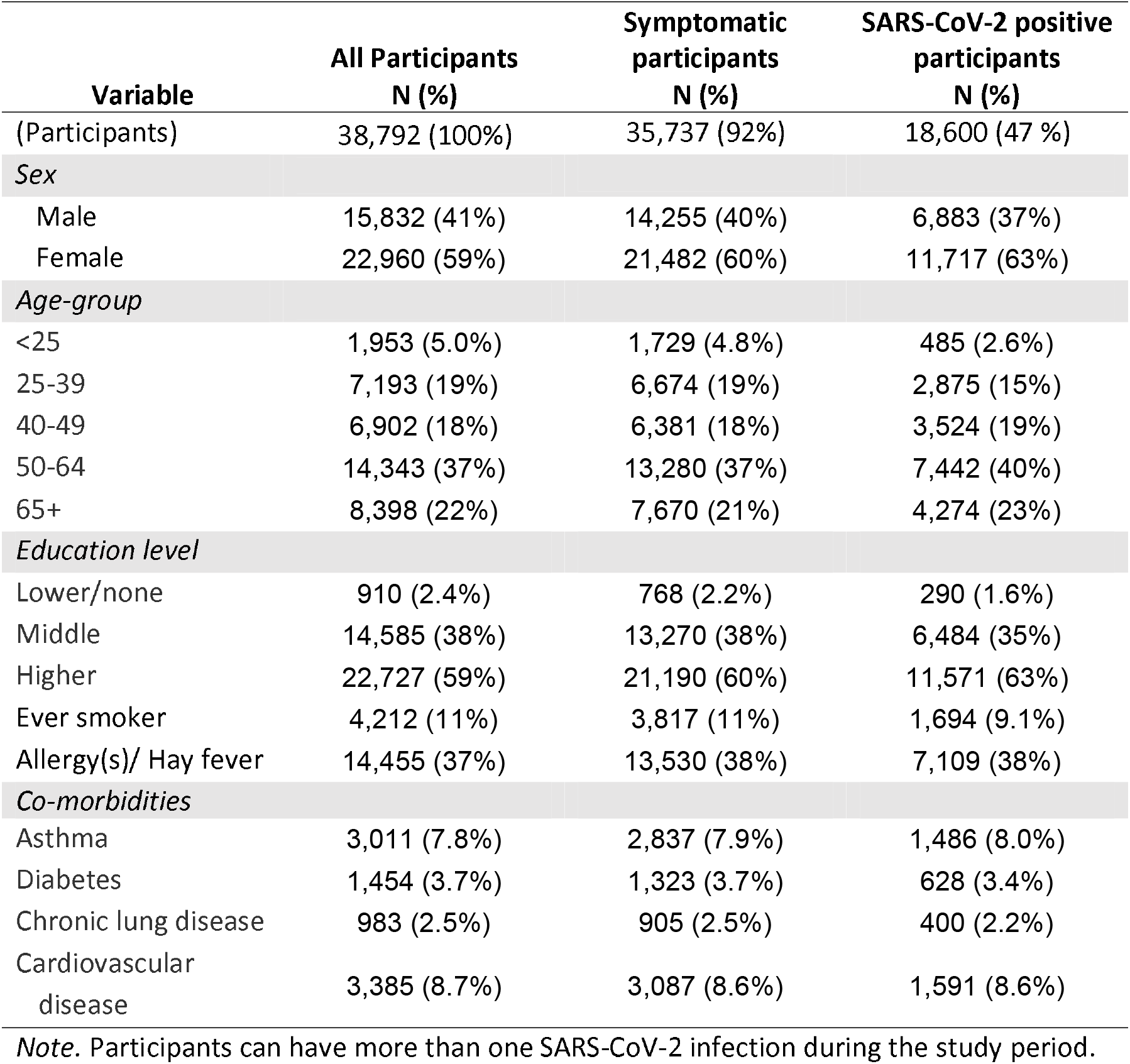
Participant characteristics.

The proportion of participants reporting a positive SARS-CoV-2 test over the study period is shown in **Figure 1A**. In November 2020, used as the reference month in subsequent analyses, the percentage of participants reporting positive tests ranged from 0.4% to 0.8%, likely reflecting infections caused by ancestral (wild type) variants of SARS-CoV-2, before the emergence of the Alpha variant and subsequent variants of concern (**Supplemental Figure S1**). Peaks in reported positive tests occurred in March 2022 (6%) during the circulation of Omicron BA.2, June 2022 (8.5%) and September 2022 (5%) when Omicron BA.5 was predominant, and in December 2023 (4%) as JN.1 became the dominant circulating variant (see **Supplemental Figure S2**).

**Figure 1.**
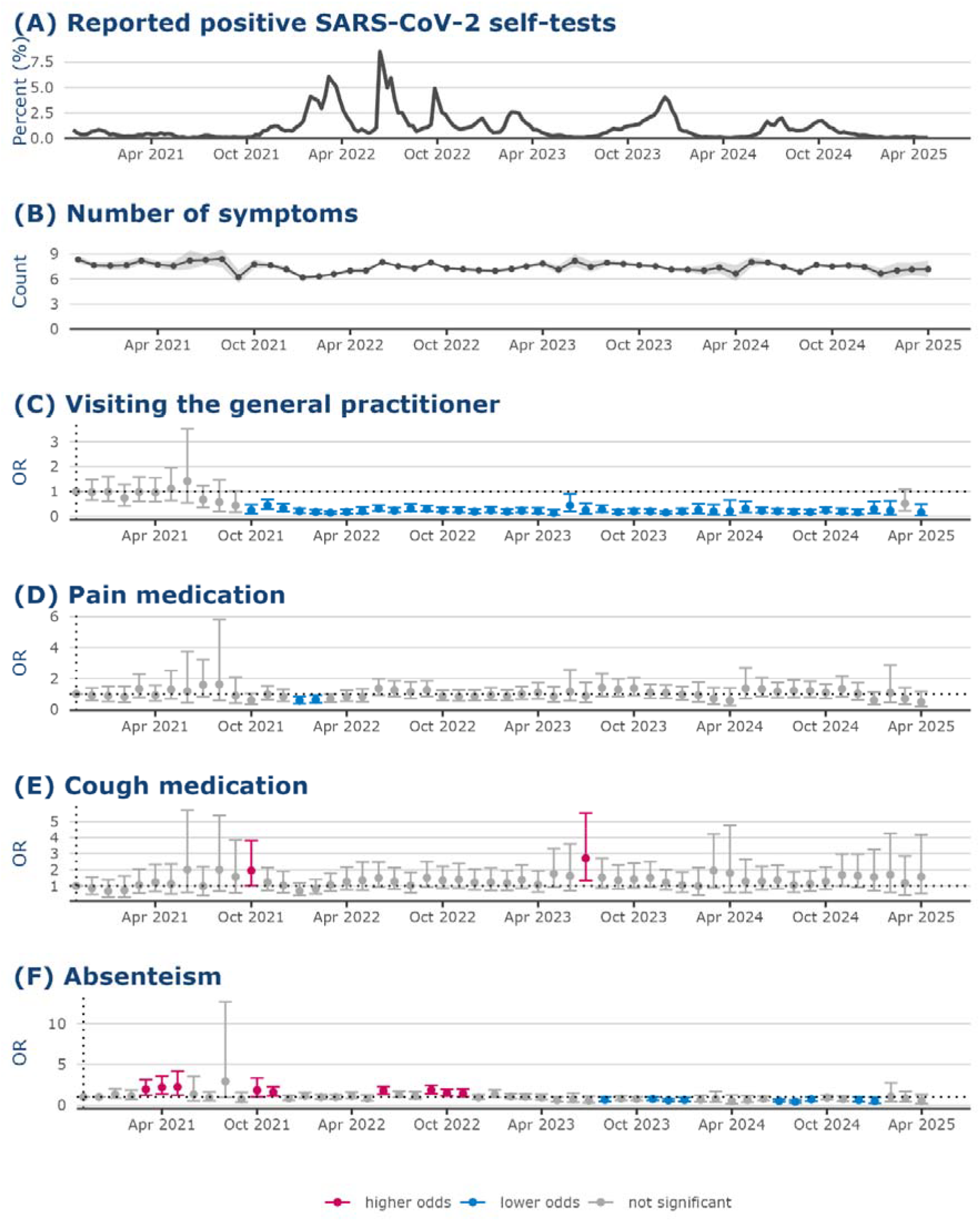
Changes in health indicators over time. (A) Reported positive SARS-CoV-2 self-tests and (B) number of symptoms, and results of logistic regression analyses showing the changes in odds of (C) GP visits, (D) pain medication usage, (E) cough medication usage and absenteeism among participants with COVID-19 between November 2020 and May 2025. November 2020 was used as reference month because this is a month with relatively low SARS-CoV-2 incidence while having sufficient data (indicated with a dotted vertical line). All analyses were adjusted for age group, sex, and presence of comorbidities. Red indicates significant higher odds ratios, blue indicates significant lower ORs and grey indicates non-significant odds ratios.

### Health indicators

The number of symptoms reported during a SARS-CoV-2 infection was generally around seven (**Figure 1B**). No consistent trend was observed in the number of symptoms experienced over time.

From October 2021 onwards, with the exception of March 2025, participants with SARS-CoV-2 infection had significantly lower odds of GP visits compared November 2020 (**Figure 1C**). This period corresponded with the predominance of the Delta variant, followed by the emergence of Omicron variants in December 2021 (**Supplemental Figure S2**).

Interestingly, around the same time, between January 2022 and March 2022, the odds of taking pain medication were significantly lower compared to November 2020 (**Figure 1D**). In almost all other months the odds of pain medication use did not significantly differ from November 2020. Similarly, the odds of taking cough medication were relatively stable over time apart from three separate months (**Figure 1E**).

The odds of taking sick leave among SARS-CoV-2 positive participants were significantly higher during March–May 2021, October–November 2021, June 2022, and September– November 2022, compared to November 2020 (**Figure 1F**). After these periods, the odds ratio (OR) for sick leave fluctuated around 1, with some months showing a significantly lower likelihood of sick leave.

### Clinical symptoms

The clinical presentation of SARS-CoV-2 infection in our study population changed over time (**Figure 2**; see **Supplemental Figure S3** for details). Compared to November 2020, the odds of reporting loss of smell and taste gradually declined, with a marked and sustained reduction after Omicron emerged in December 2021. From October 2021, the odss of rhinorrhea increased significantly, as did sneezing and sore throat from late 2021 and early 2022, respectively. Dyspnea decreased during the first year of Omicron, while cough increased after November 2020 but was no longer elevated by February–April 2025. The odds of systemic symptoms among symptomatic SARS-CoV-2 infected participants like fever and chills remained stable. Most gastrointestinal symptoms (except vomiting) and reports of general or chest pain decreased during early spring 2022 and winter 2022/2023, while headache rates stayed stable. Overall, since autumn 2021, SARS-CoV-2 symptoms have shifted toward a higher likelihood of respiratory symptoms and lower likelihood of loss of smell/taste and gastrointestinal complaints.

**Figure 2.**
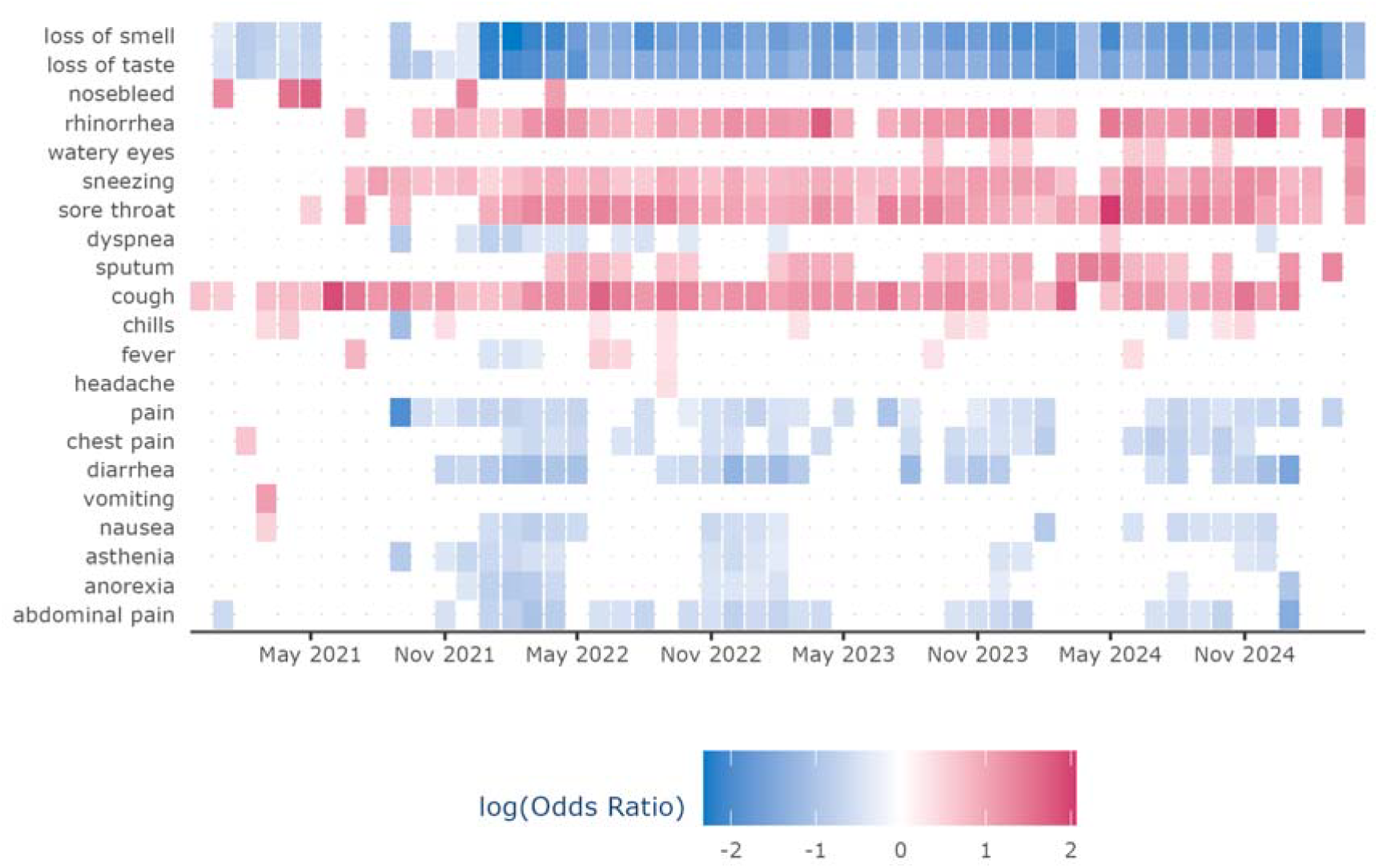
Symptoms over time. Results of logistic regression analyses showing the odds of experiencing one of the depicted symptoms among participants with COVID-19 between December 2020 and April 2025 using November 2020 as reference month. All analyses were adjusted for age group, sex and presence of comorbidities. Blue squares indicate months with significantly less reports of the relevant symptom while red squares indicate significantly higher reports of the relevant symptom compared to November 2020. White squares indicate months with non-significant odds ratios.

### Reported health score

In all age groups the marginal mean health score stayed relatively constant over time with the oldest groups constantly reporting a slightly higher health score than younger groups (**Figure 3**). This indicates that although different variants evolved over time and vaccination campaigns as well as re-infections influenced the immunity status of our participants, the overall reported burden of COVID-19 on health among those infected stayed relatively constant.

**Figure 3.**
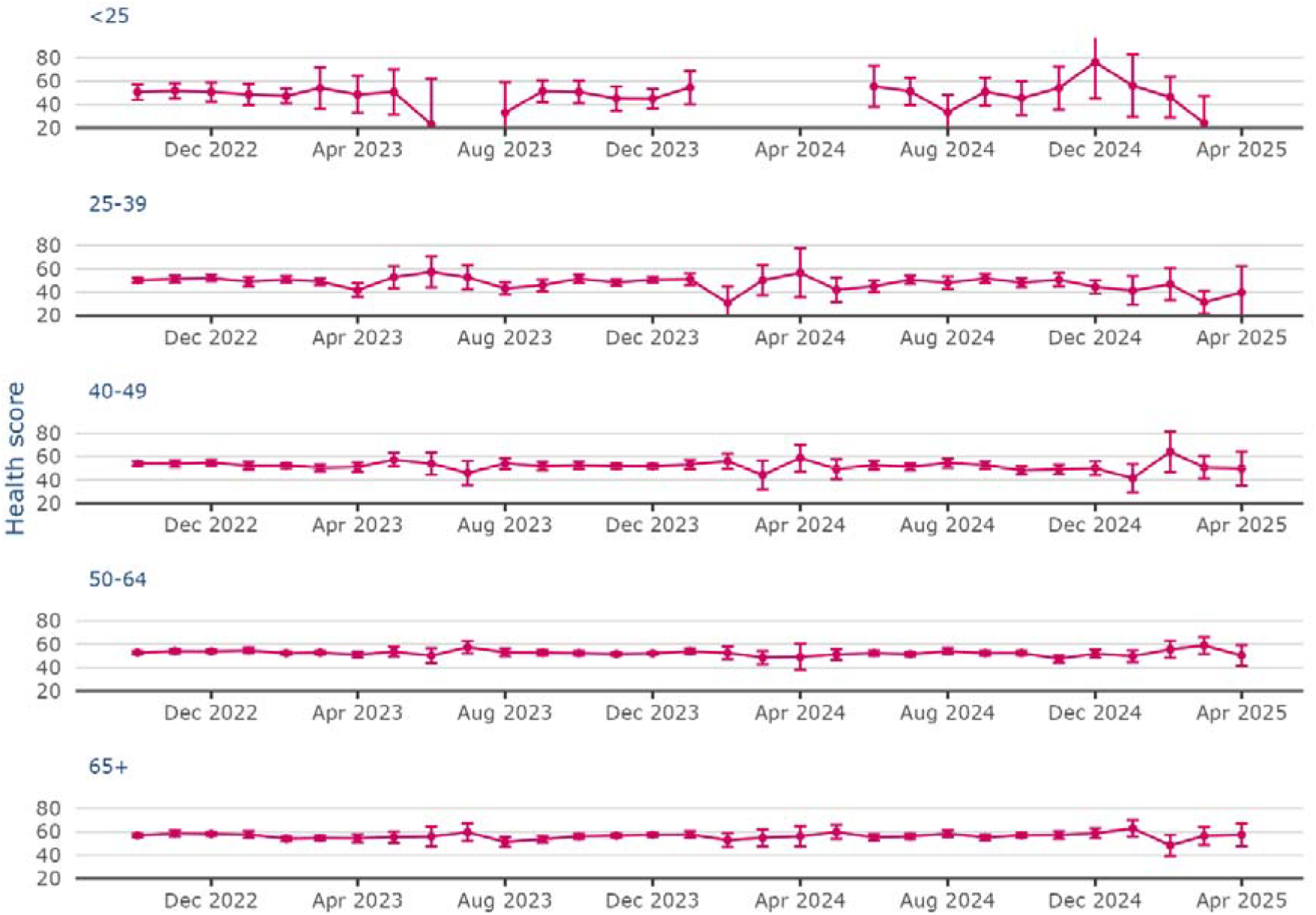
Marginal mean health score over time. Predicted mean health score of participants with COVID-19 per age group (in years) over time, adjusted for age group, presence of comorbidities and sex. Due to limited numbers of participants in the <25 year age group, it was not possible to calculate the marginal mean for certain months.

### Weighted number of symptoms and health score

We observed that those with a self-test reported a similar number of symptoms compared to a PCR-test until September 2023. From then on those with a self-test reported significantly more symptoms (**Supplemental Figure S4**). Over time, we observed a decline in testing propensity during symptomatic infections, and testing was positively associated with the number of symptoms and participation in the self-test study (**Supplemental Figure S5**). Early in the pandemic, participants thus were more likely to test for mild symptoms, potentially capturing milder cases and leading to an overestimation of severity in later periods. To assess the resulting bias on our findings, we estimated the proportion of SARS-CoV-2 cases among all symptomatic participants, stratified by number of symptoms (**Figure 4, Supplemental Part 3**). This adjustment resulted in a higher proportion of cases with few symptoms and a lower proportion with many symptoms **(Supplemental Figure S6**). The proportion of infections with fewer than five symptoms decreased until June 2021, then stabilized, while those with more than ten symptoms increased between May and September 2021 and remained stable thereafter. Periods with few positive tests, such as May–September 2023, March–May 2024, and January–April 2025, showed greater variability and wider credibility intervals.

**Figure 4.**
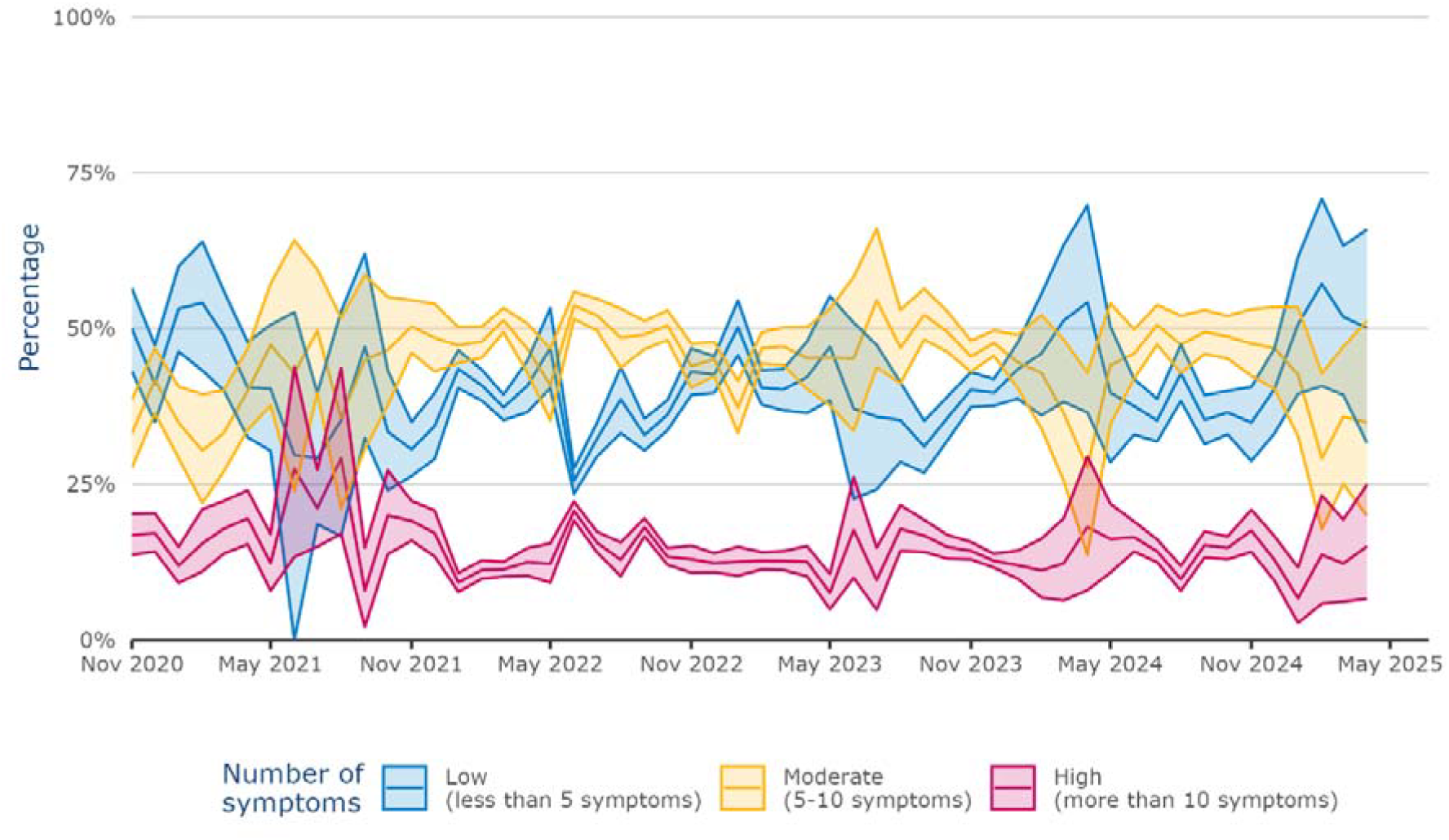
Estimated proportion of SARS-CoV-2 cases per symptom group in the symptomatic study population. All symptomatic participants (independent of the cause of their symptoms) were grouped by the number of reported symptoms during an infection episode and the SARS-CoV-2 positivity rate was used to estimate the proportion of SARS-CoV-2 cases per symptom group. Credibility intervals around these estimates were obtained by 1000 bootstrapped sampled datasets.

We repeated this analysis using self-reported health scores to calculate a weighted health score for August 2022–April 2025 (**Supplemental Figure S7**), which remained relatively constant over time. In the last three months (February–April 2025), a higher proportion of infections with high health scores and fewer with low or moderate scores were reported, with wide credibility intervals due to a low number of infections in these months. Finally, we compared participants who tested positive for SARS-CoV-2, rhinovirus, influenza A and B, and seasonal coronaviruses (NL63, HKU1, OC43, and 229E; see **Supplemental Figures S8 and S9**). About 75% of those with rhinovirus or seasonal coronaviruses reported few symptoms (less than five), compared to only 40% of those with influenza or SARS-CoV-2, suggesting that the latter two cause a higher disease burden. Similarly, participants with rhinovirus or seasonal coronaviruses consistently had higher health scores, indicating better health, than those with SARS-CoV-2 or influenza. Only 20% of SARS-CoV-2 and influenza cases had high health scores (above 65), versus 60% for rhinovirus and seasonal coronavirus cases. In summary, SARS-CoV-2 disease burden remained constant over time and was similar to that of influenza, with more severe health impact than observed with other respiratory infections.

## DISCUSSION

In this study, we examined changes in self-reported symptoms and associated burden among the general population infected with SARS-CoV-2 over a period of 4.5 years, from November 2020 to April 2025, using data from the participatory surveillance platform Infectieradar in the Netherlands. We observed a shift in type of symptoms: upper respiratory symptoms like rhinorrhea, sneezing, and sore throat became more prominent, and previously characteristic symptoms such as loss of smell and taste became less prevalent. We did observe a reduction in general practitioner consultations among those with a positive test, from October 2021 onwards. Importantly, the number of symptoms and the self-reported health score remained relatively stable over time and were more comparable with influenza virus infections than with other respiratory infections.

A notable symptom in the pre-Omicron era, particularly with the Delta variant, was the loss of smell and taste. This symptom has been potentially attributed to a mutation in the SARS-CoV-2 spike protein, which enhanced the virus’s binding affinity in the olfactory epithelium and taste buds (13). The emergence of early Omicron variants in 2021 marked significant changes in the evolution of SARS-CoV-2. Omicron BA.1 had numerous mutations compared to Delta, while a shift in the clinical presentation of the infection was observed, with Omicron variants causing more commonly sore throat, increased rhinorrhea and sputum production, while loss of smell and taste were reduced (14–16). This is in line with our results showing that upper respiratory symptoms became more prominent along with the reduction of loss of smell and taste during mild infections in the general population.

These changes in symptom profiles, combined with evolving public attitudes and pandemic restrictions, had an impact on healthcare-seeking behavior. Like many countries, the Netherlands experienced significant disruptions to routine healthcare during the COVID-19 pandemic, with only non-deferrable appointments allowed during the first wave. Additionally, many individuals may have avoided visiting their GP or hospital (17). A study in the Netherlands found that only one third of people with COVID-19-like symptoms visited their GP during the first wave of the pandemic (18). Healthcare avoidance was linked to poor self-rated health, anxiety, and fear of infection, not feeling ill enough and worries about straining the already burdened healthcare system (19,20). In later stages of the pandemic, reduced GP visits among individuals infected with SARS-CoV-2 may have reflected a shift in disease perception, with patients potentially being less concerned about their symptoms and increasingly accustomed to managing them through self-care. These factors together could explain why we observed fewer GP visits from October 2021 onwards.

Although we would expect disease severity to decline over time as a result of widespread vaccination and repeated infections, our study observed a similar pandemic and endemic symptomatic disease burden. This unexpected finding may be explained by the characteristics of vaccine-induced immunity, which is highly effective at preventing severe disease but provides only limited and short-lived protection in the upper respiratory tract, where SARS-CoV-2 first establishes infection (21,22). While natural infection induces stronger local mucosal (IgA) responses, these also decline rapidly, leaving individuals — even those with hybrid immunity gained through both vaccination and prior infection — susceptible to reinfection, especially with immune-evasive variants (24, 25). However, prior immunity still protects well against severe outcomes, as breakthrough and reinfection cases rarely lead to hospitalization or death in vaccinated individuals (26,27).

Given these complex interactions of the immune system and SARS-CoV-2, we opted against a variant-specific analyses. Although specific SARS-CoV-2 variant information of part of our study population was available, without detailed immune profiling of our participants and information on vaccination status or prior infections, attributing changes in symptom burden to specific variants alone would oversimplify these interactions. Also, our goal was to assess trends in symptomatology and its perceived burden over time, not per variant.

Our study had several strengths, including the large cohort of individuals, real-time data on symptoms and self-reported health during SARS-CoV-2 infection not selected on health care seeking behavior. However, limitations include an overrepresentation of older, female, and highly educated participants, which may affect generalizability. Moreover, the health status used to determine the severity of the infection reflects the individually perceived health, which is inherently subjective. Despite observing consistent trends in health scores over time, caution is necessary when making direct comparisons about disease severity between participants. Additionally, we rely on participants to voluntarily perform a SAR-CoV-2 self-test when experiencing symptoms. Availability of self-tests before the start of the self-swab study and willingness to perform a test especially towards the end of the pandemic and beyond might have led to an increasing overestimation of SARS-CoV-2 symptom burden even though we tried to correct for this.

Overall, our findings suggest that there are changes in clinical presentation of COVID-19 with symptoms developing towards upper respiratory tract associated symptoms. However, the endemic symptomatic disease burden of COVID-19 in the general population based on the number of symptoms and self-reported health score stayed constant over time, being comparable to influenza virus infections. This underscores the ongoing disease burden and public health relevance of SARS-CoV-2, even as population immunity has increased and the risk of severe outcomes like hospitalizations has declined.

## Data Availability

Not all data can be shared publicly because of legal constrains. Data underlying the results are also available on www.infectieradar.nl/results and data requests can be send to.

## Funding Source

This work was supported by the Ministry of Health, Welfare and Sports (VWS), the Netherlands. The funders had no role in study design, data collection and analysis, decision to publish, or preparation of the manuscript.

### Ethical approval statement

The samples were collected as part of Infectieradar. The protocol for Infectieradar was approved by the Medical Ethics Review Committee Utrecht (reference number: WAG/avd/20/008757; protocol 20-131) was obtained given the nature of data collection. All participants provided informed consent upon registration, which included agreement to the privacy statement and described the processing of personal data and research results, website security measures taken, and how to file a complaint. Participants were eligible to withdraw from the study at any time. Individuals had to be 16 years or over to be able to participate.

## Acknowledgements

We thank the participants of Infectieradar for their weekly contributions and submission of samples. Without their contribution this project would not have been a success. Furthermore, we thank our colleagues who help maintain and run the Infectieradar-cohort and handle the samples. Developing and running this study has been a huge team effort.

