## Supplemental for "Longitudinal trends in the endemic symptomatic burden of COVID-19: Insights from community-based participatory virological surveillance in the Netherlands, November 2020 - April 2025"

**SUPPLEMENT**

**Part 1: Variant determination without lab samples**

We assigned variants causing SARS-CoV-2 infections in participants before the addition of the self-testing component in Infectieradar based on the date of a self-test or PCR test. Variant periods were defined based on national surveillance data as periods in which one variant was detected in at least 90% of all tested samples with minimal co-circulation of other variants (8). The following periods were defined: Alpha (22 March to 31 May 2021), Delta (12 July 2021 to 13 December 2021), Omicron BA.1 (10 January 2022 to 24 January 2022), Omicron BA.2 (21 March 2022 to 2 May 2022) and Omicron BA.5 (18 July to 29 August 2022). No time periods were assigned to Beta and Gamma due to low percentage of samples testing positive for these variants, making them indistinguishable from other variants using the 90% threshold.

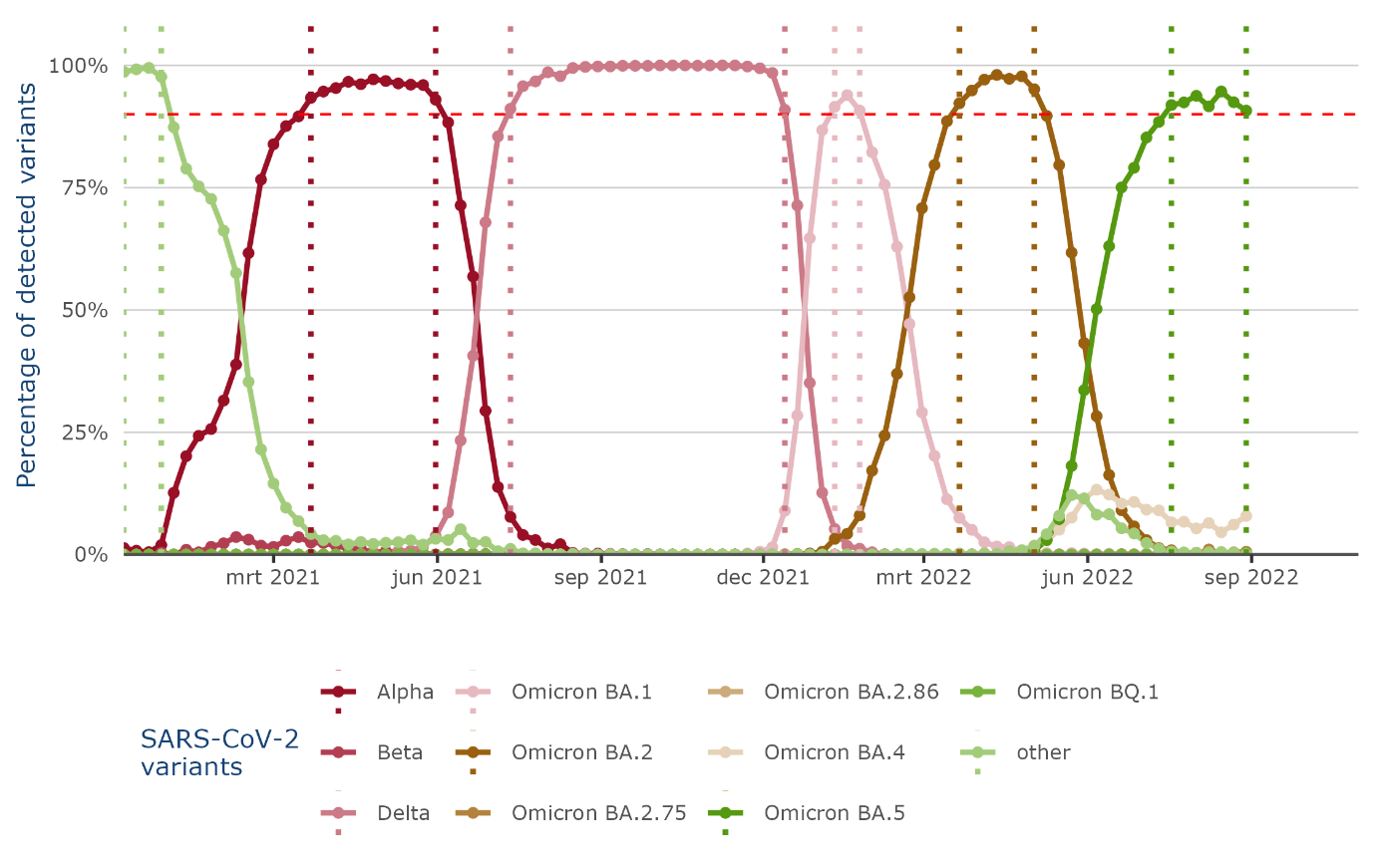
 **Figure S1: Cut-off dates used for variant detection.** Figure shows percentage of detected variants in the national surveillance of the Netherlands. We used a 90% cut-off (red dashed line) to determine periods in which one variant made off more at least 90% of all tested monsters.

Between April 2021 and September 2022 Infectieradar participants reported 2,516 infection episodes with a positive SARS-CoV-2 self-test or PCR test that could be assigned to a SARS-CoV-2 variant (Figure S2A). Of those infection periods, 5.9% (148/2516) were assigned to the Alpha variant, 24.9 % (626/2516) to the Delta variant, 12.2% (307/2516) to Omicron BA.1, 38.4% (967/2516) to Omicron BA.2 and 18.6 % (468/2516) to Omicron BA.5.

The number of NTS tested positive per week between October 2022 and May 2024 greatly changes over time with peaks in week 41 2022 and week 8 and 50 2023 with over 90 samples tested positive for SARS-CoV-2 in those weeks (Figure S2B). During the summer of 2023 from May to August the number of samples testing positive for SARS-CoV-2 per week stayed below 15 samples per week indicating less circulation of SARS-CoV-2 in our study population during this time. The most dominant SARS-CoV-2 variant per week changed over time: BA.5 and BQ.1 circulated since the start of the self-swab study until in February 2023 XBB.1.5 and XBB.1.9 took over. BA.2.86 was detected at low proportions since the start of the study until June 2023. At the end of the summer 2023, when the number of samples per week were increasing again, EG.5 started to circulate but was outcompeted by JN.1 making up the largest proportion of all positive samples from November 2023 to the end of May. Then, the most recent variants, KP.2 and KP.3 dominated until the end of the study period.

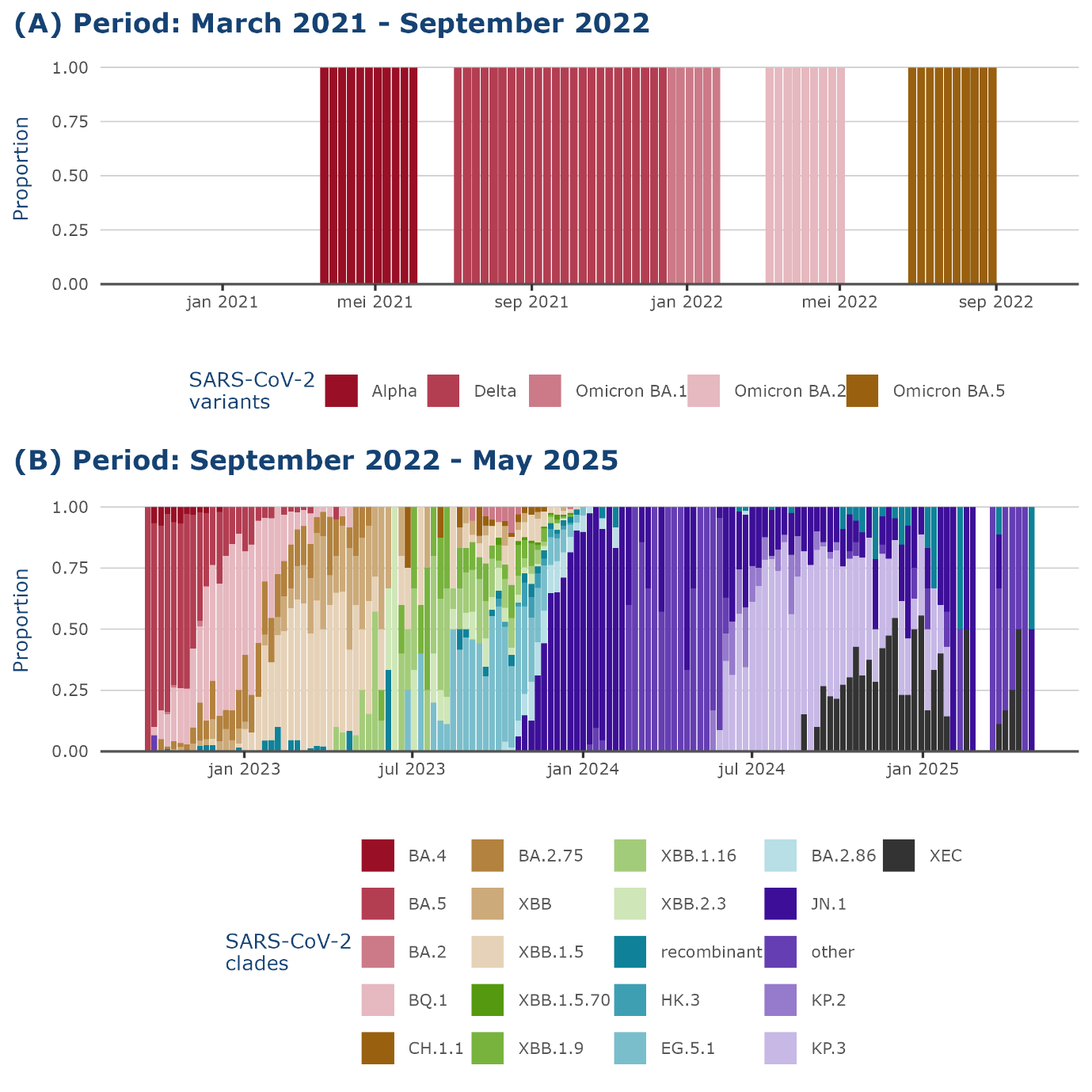

**Figure S2. Reported SARS-CoV-2 variants over time.** The self-swab study of Infectieradar was added in September 2022. Before that time, reported SARS-CoV-2 positive tests where assigned to a SARS-CoV-2 variant based on the reported date of the positive PCR or self-test (A). After September 2022 the self-swab study was started and SARS-CoV-2 positive nose- and throat samples were sequenced to determine the SARS-CoV-2 variant (B). *Note*: in week 15 in 2024 no SARS-CoV-2 positive samples were detected.

**Part 2: Additional figures**

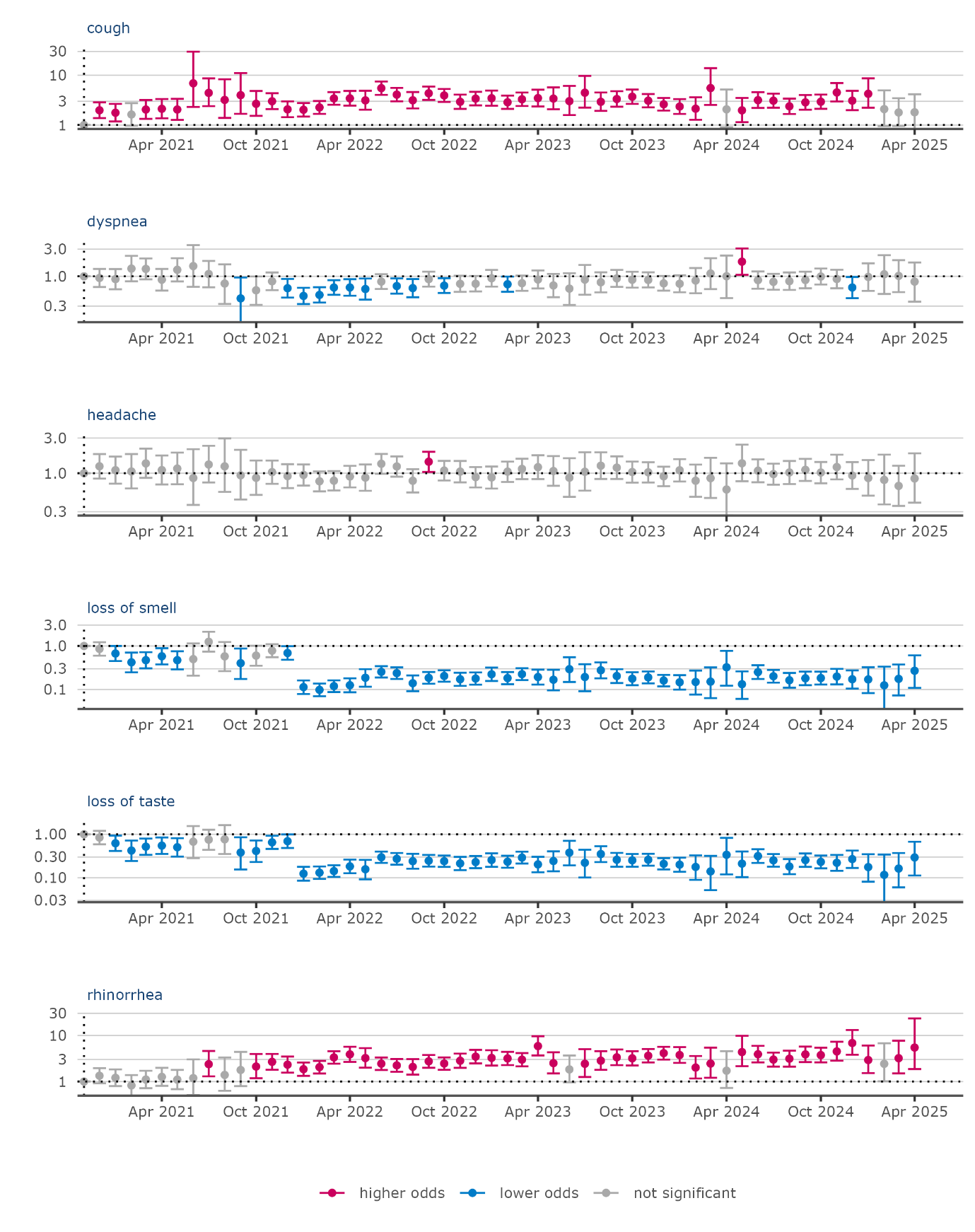

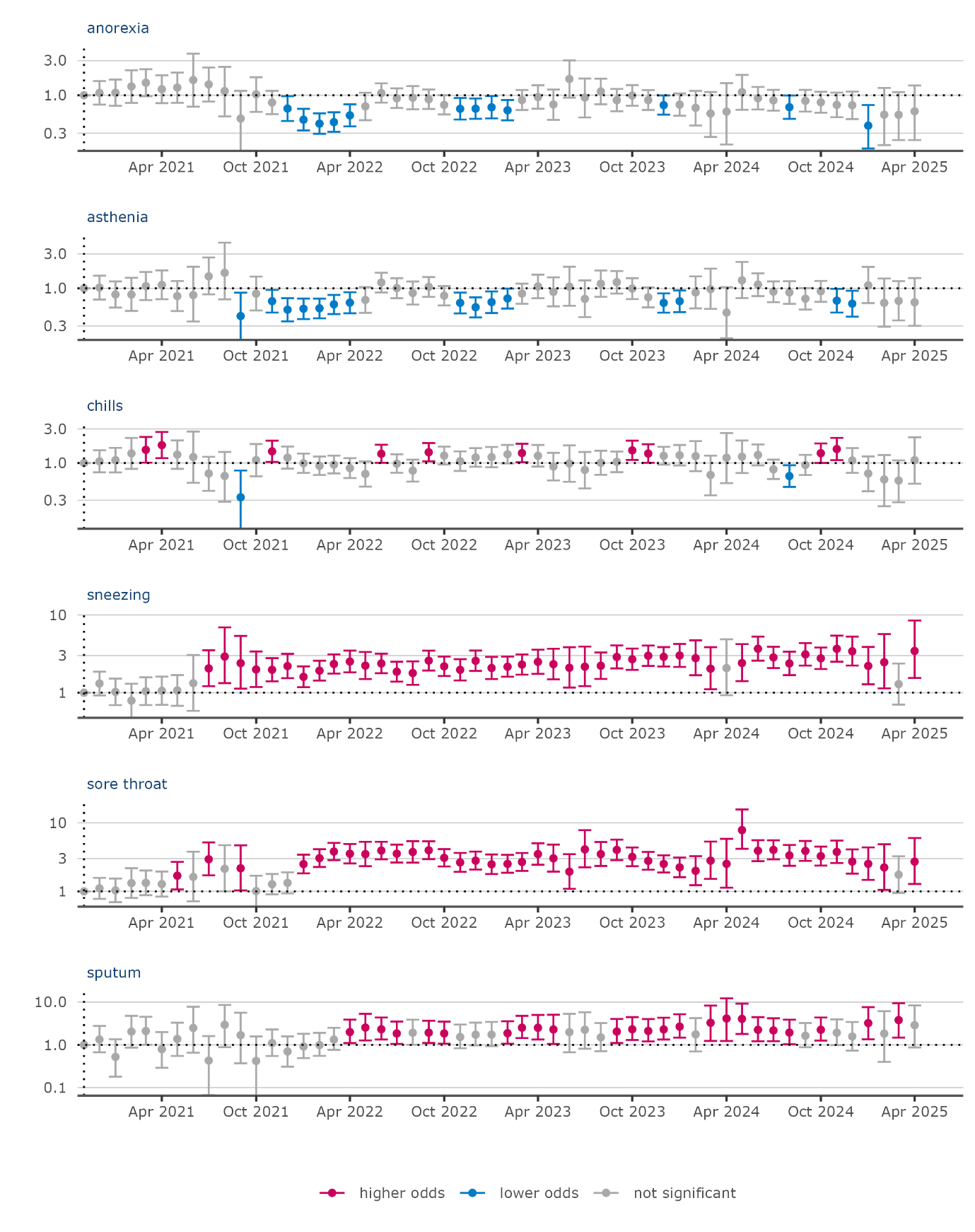

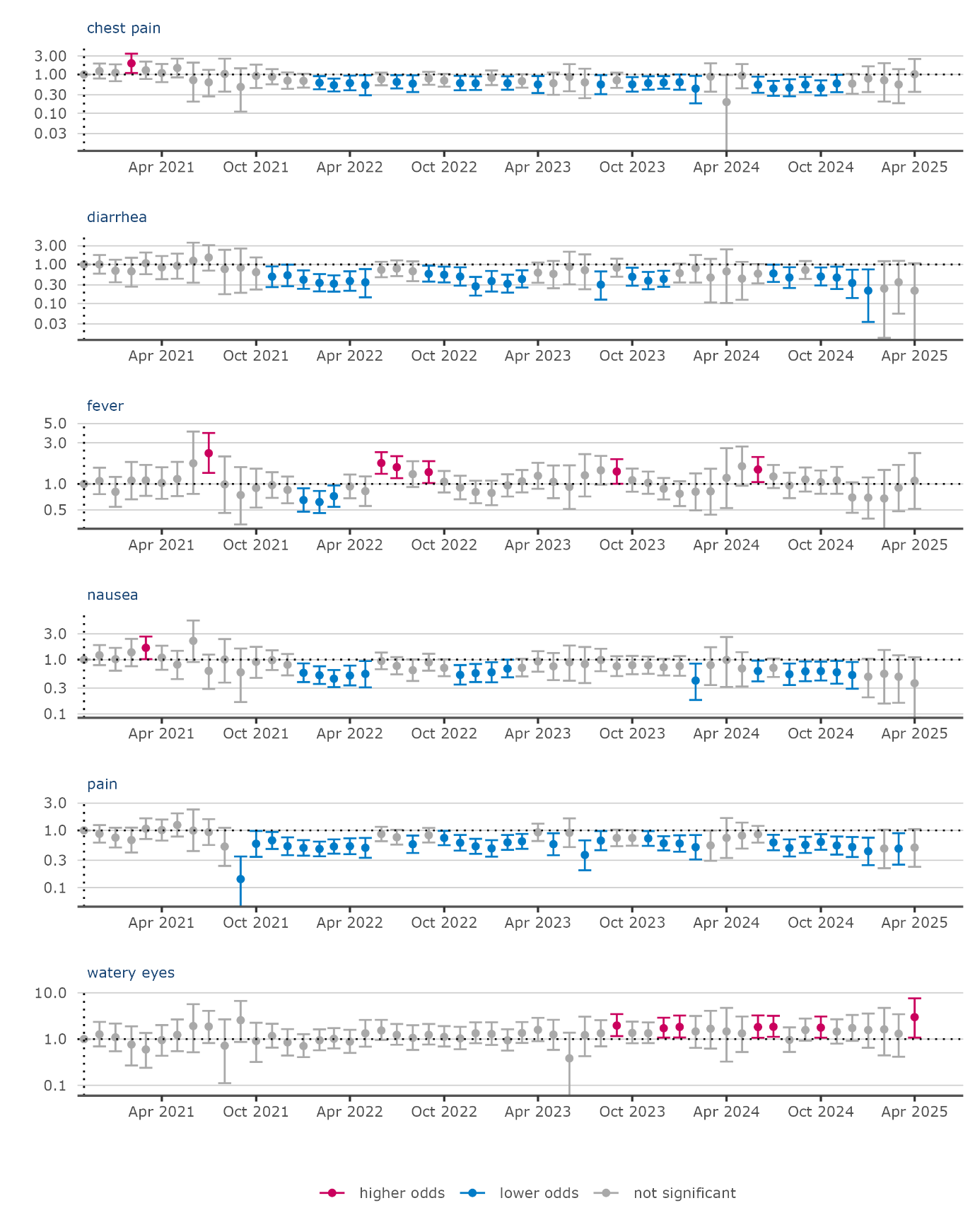

**Figure S3. Adjusted odds ratios of experiencing symptoms per month**. Results of regression analyses showing the odds of experiencing one of the depicted symptoms for participants with COVID-19 between November 2020 and April 2025. November 2020 was used as reference month because this is a month with relatively stable SARS-CoV-2 incidence while having sufficient data (indicated with a dotted vertical line). All analyses were adjusted for age group, sex and presence of comorbidities.

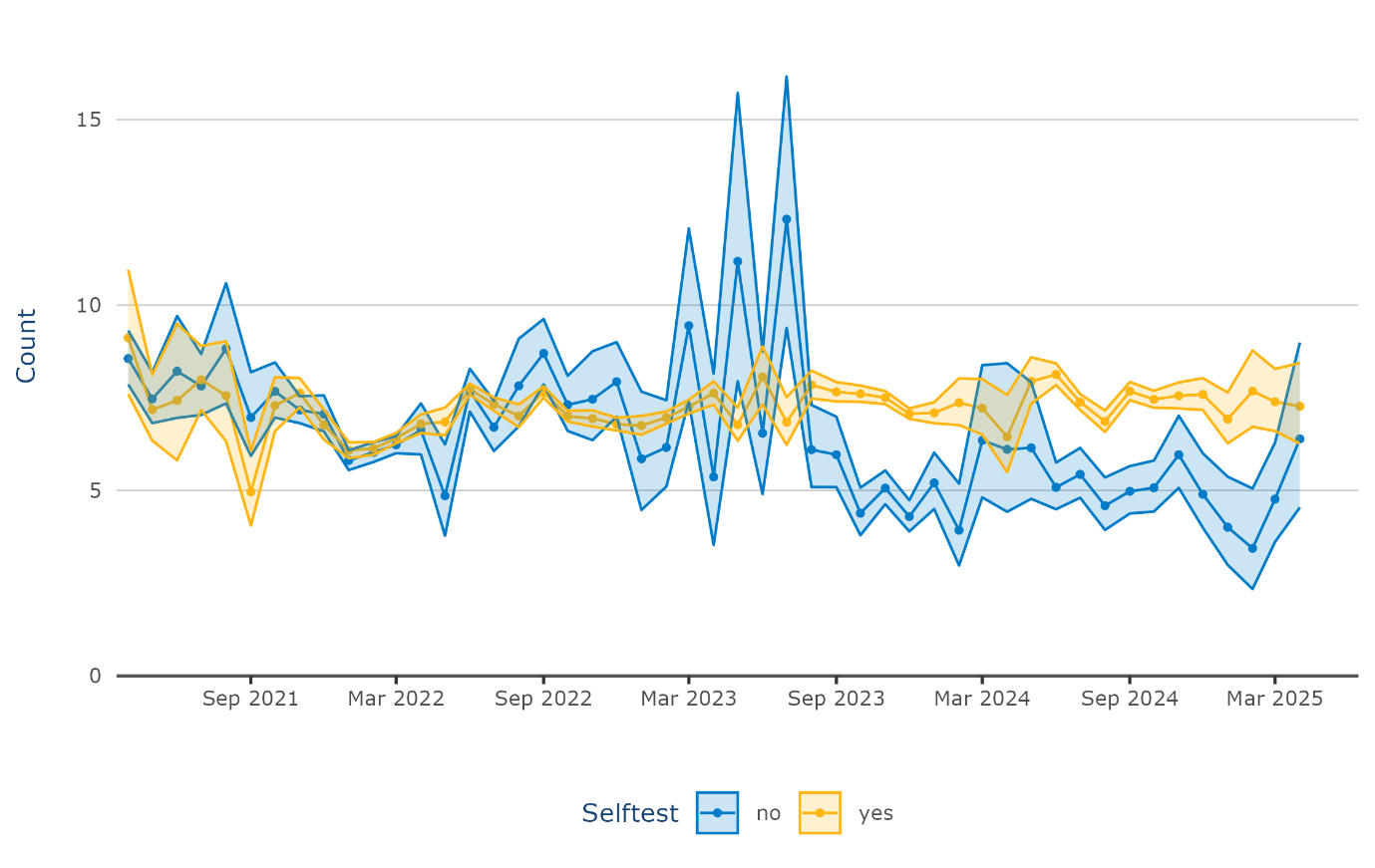

**Figure S4: Difference in number of reported symptoms.** Marginal means of reported symptoms by participants with positive self-test and participants with PCR-test only. Analyses were adjusted for age group, sex and presence of comorbidities.

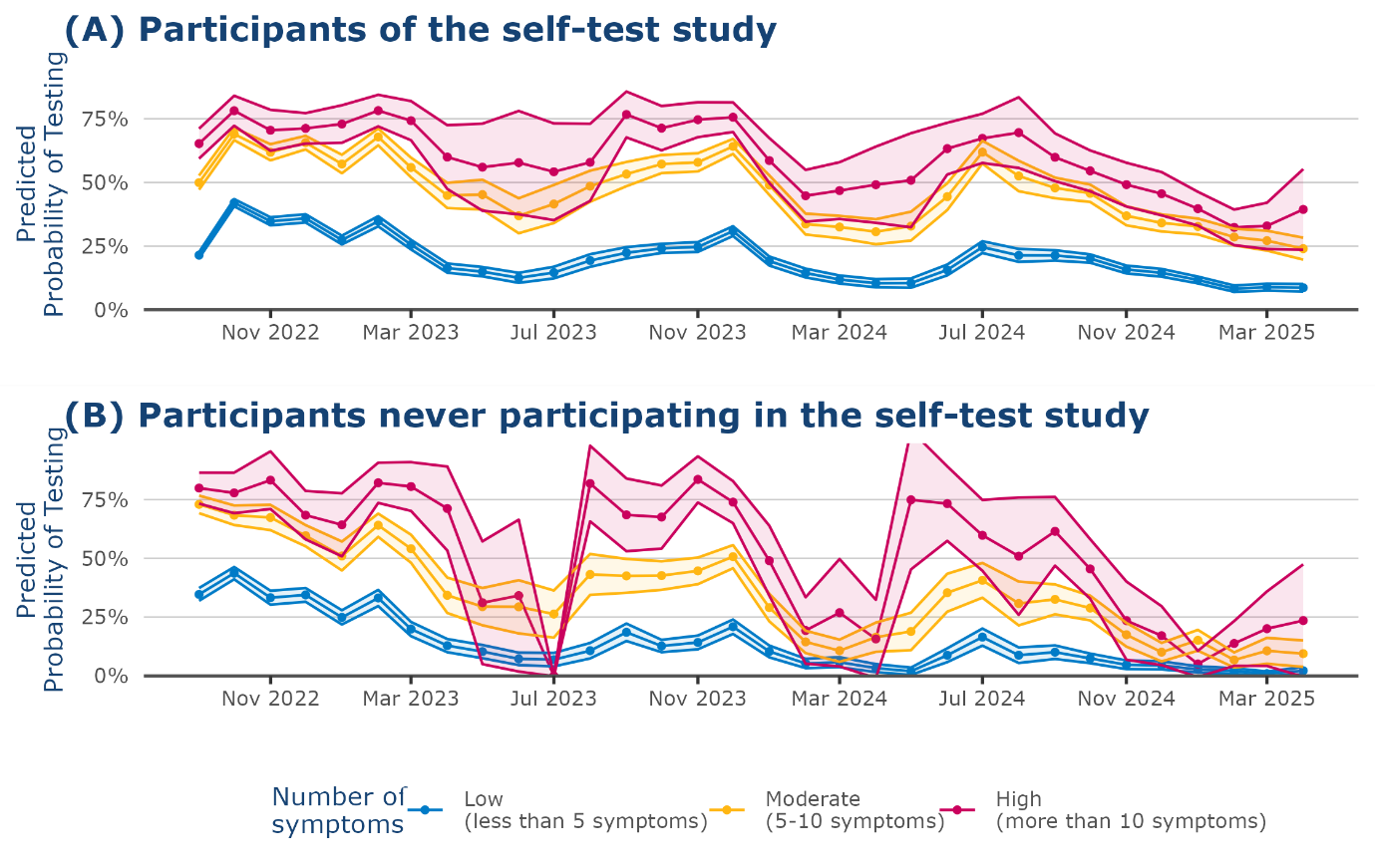

**Figure S5: Changes in testing behavior over time.** Predicted probability to perform a SARS-CoV-2 self-test stratified by the number of symptoms per month between September 2022 and May 2025 for (A) active participants of the self-swab study (n = 13,013; participants participating in the self-swab study at the time of the test) and (B) participants who never participated in the self-swab study of Infectieradar (n = 6,479). Participants, who entered the self-swab study but quit during the study period were excluded from this overview (n = 613). *Note*: The self-swab study started in September 2022.

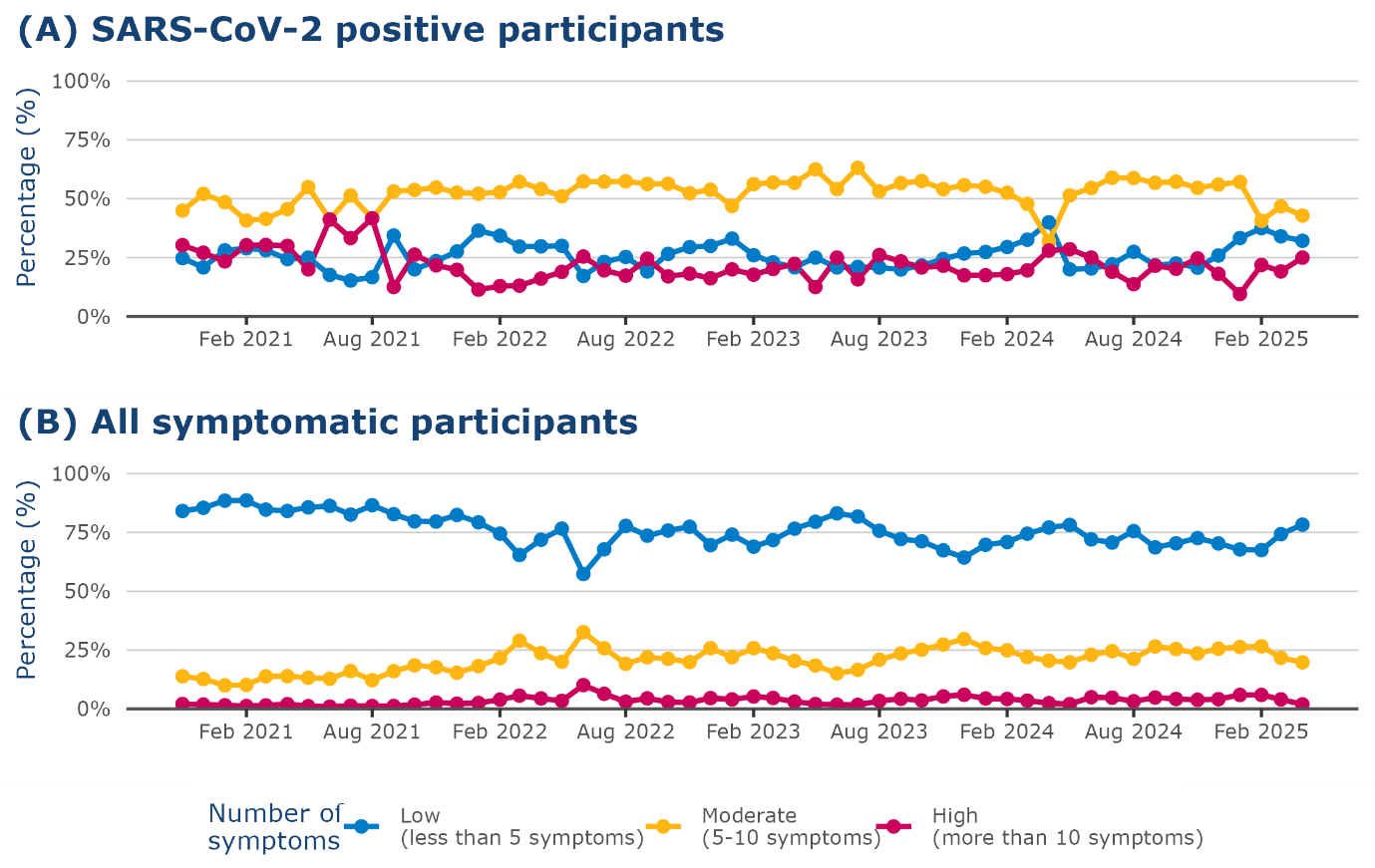

**Figure S6 Distribution of number of symptoms per week.** Distribution of reported number of symptoms among (A) SARS-COV-2 positive participants and (B) symptomatic participants (independent of the cause of their infection) per week.

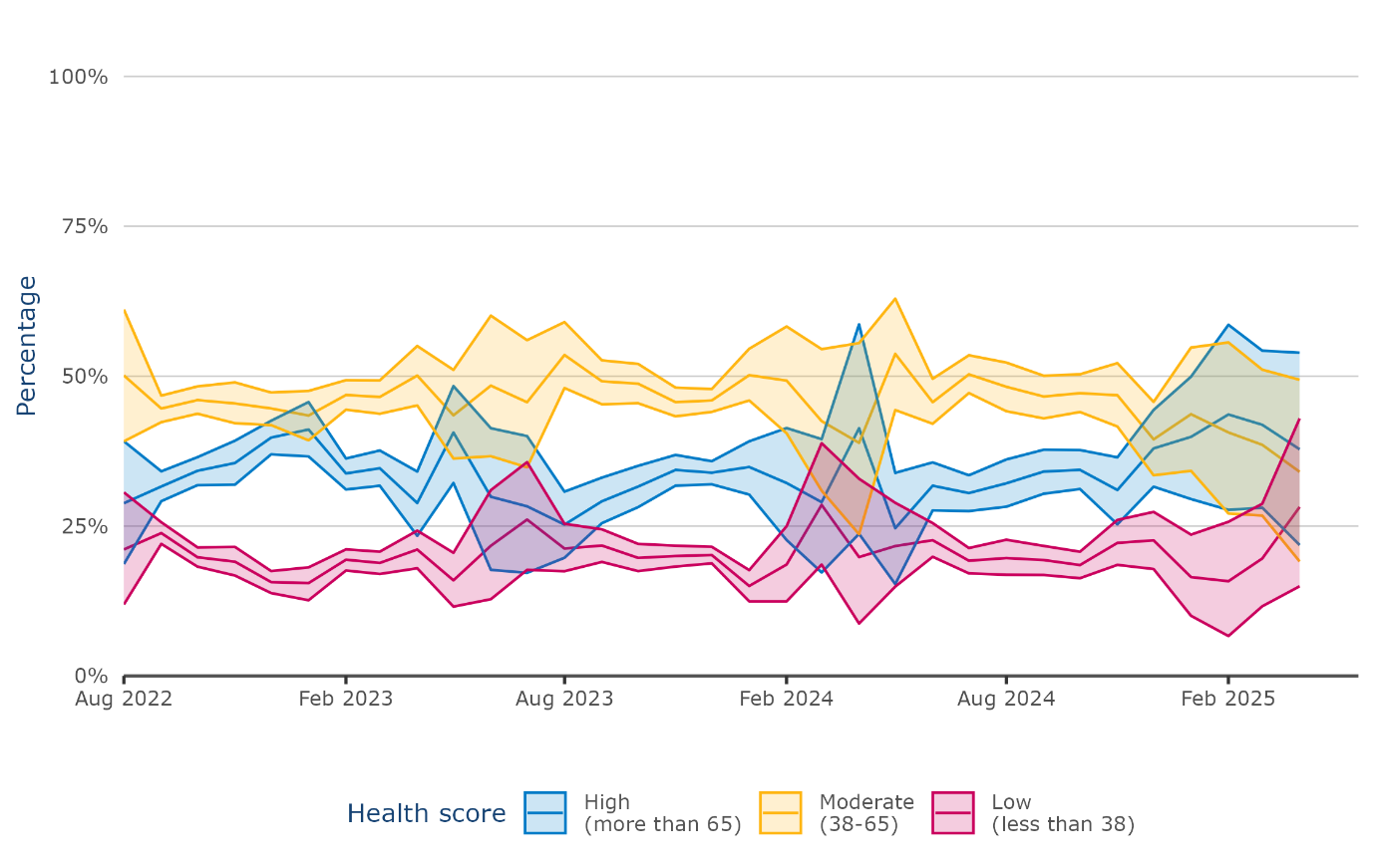

**Figure S7** **Estimated proportion of SARS-CoV-2 cases per health score group in the symptomatic study population.** All symptomatic participants (independent of the cause of their symptoms) were grouped by their reported health score (from 0 – 100, with 0 being the worst possible health, and 100 being the best possible health) during an infection episode and the SARS-CoV-2 positivity rate was used to estimate the proportion of SARS-CoV-2 cases per health score group. Credibility intervals around these estimates were obtained by 1000 bootstrapped sampled datasets. Data for this analysis was available from August 2022 – April 2025.

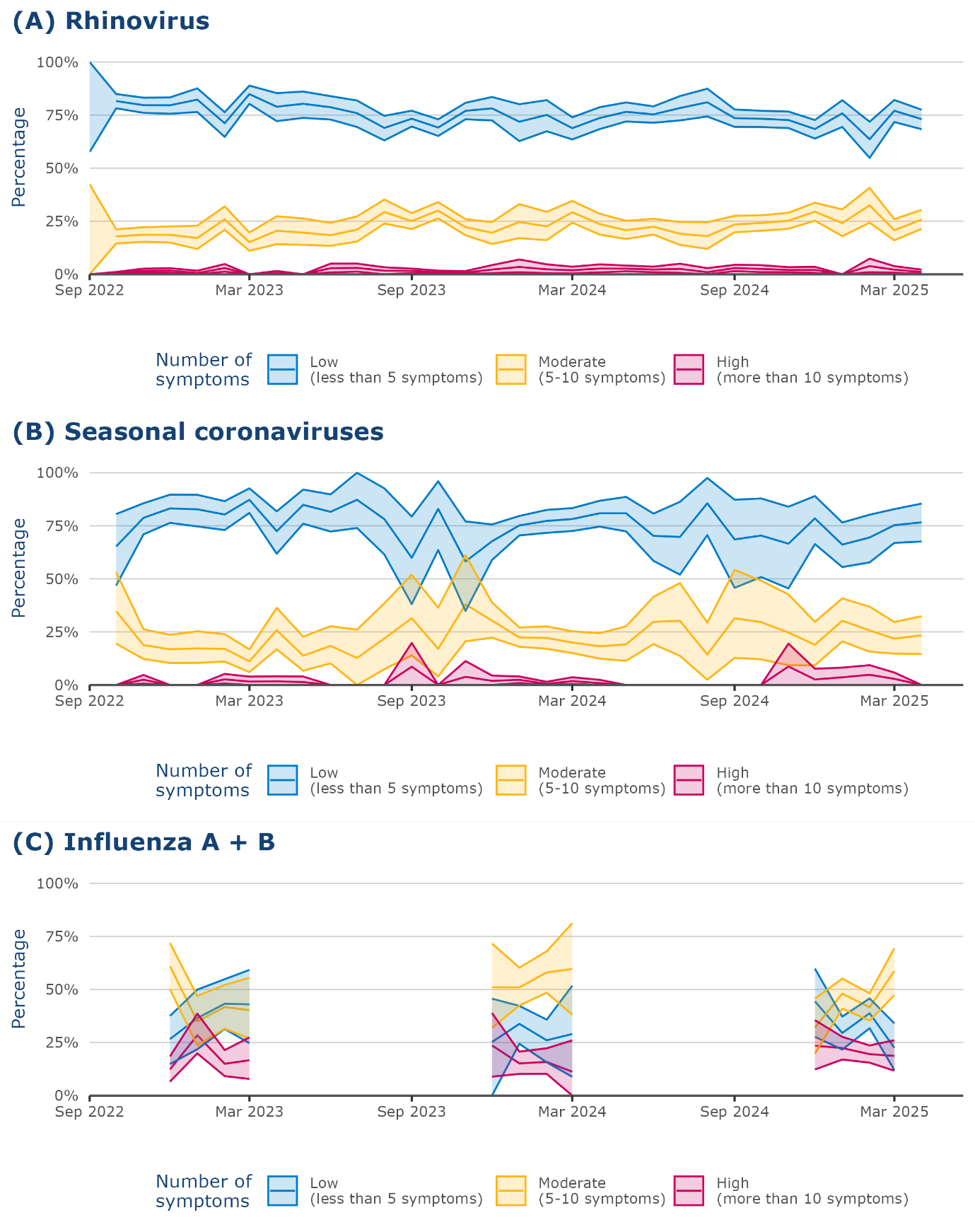

**Figure S8** **Estimated proportion of cases per pathogen per symptom group in the symptomatic study population.** All symptomatic participants (independent of the cause of their symptoms) were grouped by the number of reported symptoms during an infection episode. Cut-off values for these groups were based on the 25^th^ and 75^th^ percentiles of the reported number of symptoms of the SARS-CoV-2 positive participants to allow comparison with the SARS-CoV-2 specific estimates. The positivity rate per pathogen based on the submitted nose- and throat samples from the self-swab study was used to estimate the proportion of cases per pathogen per symptom group. Credibility intervals around these estimates were obtained by 1000 bootstrapped sampled datasets. Due to the absence of positive Influenza A and B samples in certain months, these were removed from the analysis.

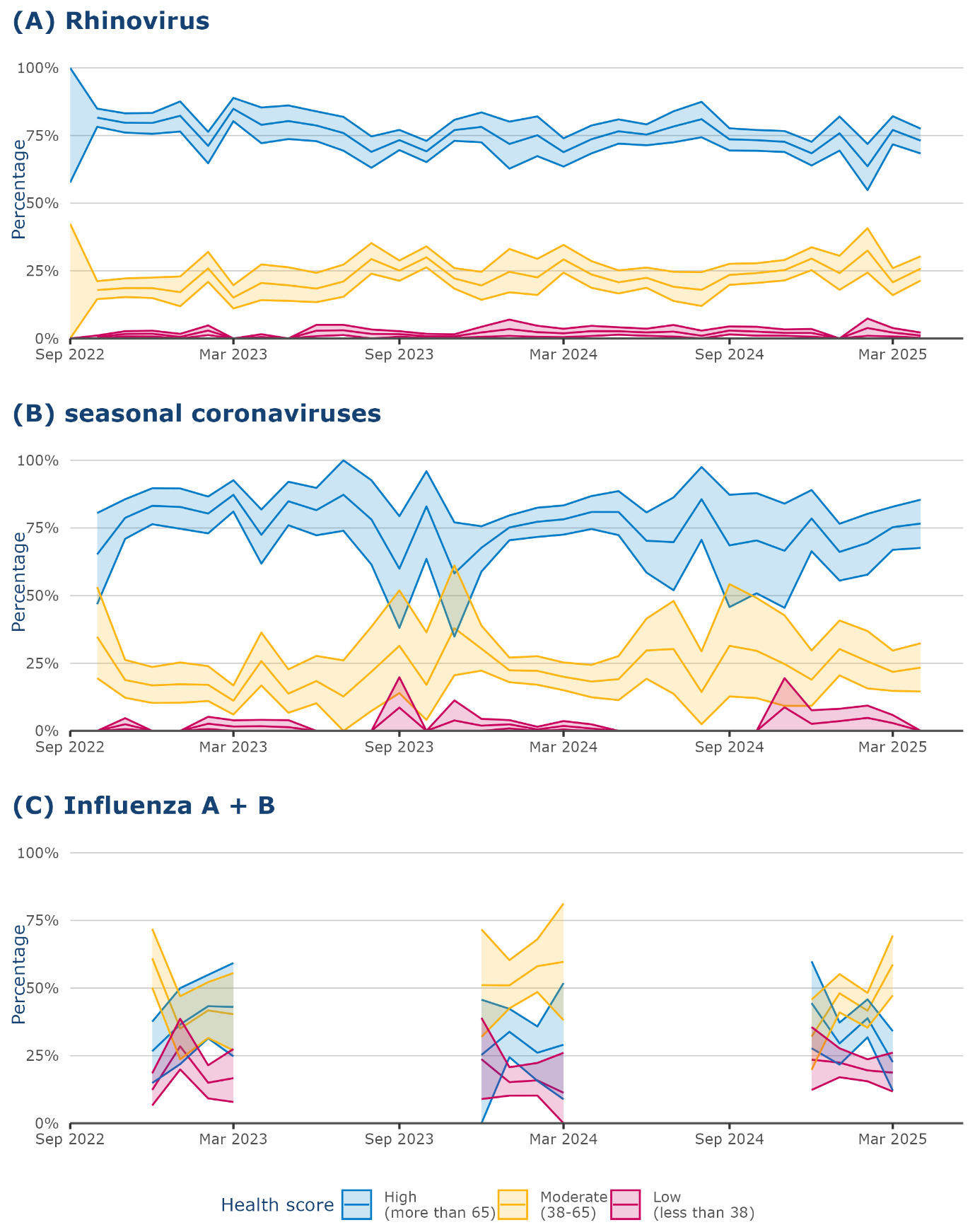

**Figure S9 Estimated proportion of cases per pathogen per health score group in the symptomatic study population.** All symptomatic participants (independent of the cause of their symptoms) were grouped by their reported health score (from 0 – 100, with 0 being the worst possible health, and 100 being the best possible health) during an infection episode. Cut-off values for these groups were based on the 25th and 75th percentiles of the reported lowest health score of the SARS-CoV-2 positive participants to allow comparison with the SARS-CoV-2 specific estimates. The positivity rate per pathogen based on the submitted nose- and throat samples from the self-swab study was used to estimate the proportion of cases per pathogen per health score group. Credibility intervals around these estimates were obtained by 1000 bootstrapped sampled datasets. Due to the absence of positive Influenza A and B samples in certain months, these were removed from the analysis. Data for this analysis was available from August 2022 – April 2025.

**Part 3: Example calculations for the weighted disease burden**

The inclusion of our SARS-CoV-2-positive study population depends on our participants’ willingness to perform a SARS-CoV-2 self-test or nose- and throat swab (NTS) when experiencing symptoms. This introduces potential bias, particularly if testing behavior changes over time dependent on the symptom burden. To assess this, we calculated a weighted disease burden taking into account the symptom burden in symptomatic participants independent of the cause of infection.

For this example we assume that we have 300 participants of a hypothetical population who report symptoms in a certain week. During this week SARS-CoV-2, influenza virus and rhinovirus circulate and could cause the symptoms. We further assume that rhinovirus mostly causes a low number of symptoms while influenza causes mostly infections with a high number of symptoms. We divide the participants based on their symptoms in three groups: low (< 2 symptoms), moderate (2-4 symptoms), high (> 4 symptoms). In each group there are 100 participants.

Now we assume two scenarios: (1) In the first scenario the probability to perform a SARS-CoV-2 self-test is independent of the number of symptoms; (2) in the second scenario participants with more symptoms have a higher probability to perform a self-test.

**Scenario 1:**

In **scenario 1**, we assume that half of the symptomatic participants in each symptom group decided to perform a SARS-CoV-2 self-test. In the group with a low number of symptoms half of those who tested (n = 50) tested positive (n = 25) for SARS-CoV-2. The positivity rate for the self-tests in this group is therefore 50% (25/50). In the moderate symptom group all 50 who tested were positive, so the positivity rate is 100%. In the high symptom group 50 participants also tested, but only 25 were positive, giving a positivity rate of 50%. The proportion of participants with a high number of symptoms among the SARS-CoV-2 positive participants is therefore 25/100 = 25%.

To find out what proportion of all positive cases had a high number of symptoms, we summed up the positives (25 in low, 50 in moderate, and 25 in high symptom group) for a total of 100 positive cases. Of these, the high symptom group contributed 25, so 25% of the positive cases were from the high symptom group.

We compare this with the weighted positivity, which takes into account the size of each symptom group. For each group, we multiply the positivity rate by the proportion of all participants in that group. For example, if the low symptom group makes up 33% of all symptomatic participants, their weighted positivity would be 50% x 33% = 17%. This tells us that 17% of all participants had both a low number of symptoms and tested positive for SARS-CoV-2.

To check the proportion of positive cases with a high number of symptoms, we add the weighted positives across all groups (for example: 17% for low, 33% for moderate, and 17% for high symptoms) and then calculate the share for the high symptom group. In this example, the resulting proportion is 25%, which matches the unweighted proportion.

This shows that if the likelihood of self-testing is the same regardless of the number of symptoms, the unweighted proportion of participants with more severe symptoms among those who test positive accurately reflects the true distribution.

**Scenario 2:**

In **scenario 2**, we consider what happens if the willingness to test depends on the number of symptoms. Here, we assume that only half of the participants with a low number of symptoms decide to take a self-test, while in the moderate and high symptom groups, all symptomatic participants get tested.

With this approach, when we look at everyone who tested positive for SARS-CoV-2, 50 out of 175 came from the high symptom group. This means that 29% of positive cases were from the high symptom group (50/175).

However, when we repeat the weighted positivity calculation—again multiplying each group’s positivity by its share of the total population—the proportion of all SARS-CoV-2 positive participants with a high number of symptoms is only 25%. This is lower than the 29% we found without weighting.

This example shows that if people with more symptoms are more likely to test, the simple (unweighted) percentage overestimates how common high symptom burden actually is among all SARS-CoV-2 infections in the population.

**Scenario 1:**

| Number of Symptoms | Symptomatic participants (n) | Proportion of symptomatic participants (%) | SARS-CoV-2 self-test (n) | | Not tested (n) | Self-test positivity (%) | Weighted positivity (%) | Proportion per symptom group (%) | |
| --- | --- | --- | --- | --- | --- | --- | --- | --- | --- |
|  |  |  | Positive | Negative |  |  |  | unweighted | weighted |
| Low | 100 | 33% | 25 | 25 | 50 | 50% | 17% | 25 % | 25% |
| Moderate | 100 | 33% | 50 | 0 | 50 | 100% | 33% | 50% | 50% |
| High | 100 | 33% | 25 | 25 | 50 | 50% | 17% | 25% | 25% |
| **Total** | **300** | **100%** | **100** | **50** | **150** |  |  |  |  |

**Scenario 2:**

| Number of Symptoms | Symptomatic participants (n) | Proportion of symptomatic participants (%) | SARS-CoV-2 self-test (n) | | Not tested (n) | Self-test positivity (%) | Weighted positivity (%) | Proportion per symptom group (%) | |
| --- | --- | --- | --- | --- | --- | --- | --- | --- | --- |
|  |  |  | Positive | Negative |  |  |  | unweighted | weighted |
| Low | 100 | 33% | 25 | 25 | 50 | 50% | 17% | 14.3% | 25% |
| Moderate | 100 | 33% | 100 | 0 | 0 | 100% | 33% | 57% | 50% |
| High | 100 | 33% | 50 | 50 | 0 | 50% | 17% | 29% | 25% |
| **Total** | **300** | **100%** | **175** | **75** | **50** |  |  |  |  |
